# A National Multicenter Evaluation of the Clinical Utility of Optical Genome Mapping for Assessment of Genomic Aberrations in Acute Myeloid Leukemia

**DOI:** 10.1101/2020.11.07.20227728

**Authors:** Brynn Levy, Linda B. Baughn, Scott Chartrand, Brandon LaBarge, David Claxton, Alan Lennon, Yassmine Akkari, Claudia Cujar, Ravindra Kolhe, Kate Kroeger, Beth Pitel, Nikhil Sahajpal, Malini Sathanoori, George Vlad, Lijun Zhang, Min Fang, Rashmi Kanagal-Shamanna, James Broach

## Abstract

Detection of hallmark genomic aberrations in acute myeloid leukemia (AML) is essential for prognosis and patient management. Clinical practice guidelines for identifying such structural variants (SVs), established by the World Health Organization (WHO), European Leukemia Net (ELN) and National Comprehensive Cancer Network (NCCN), rely substantially on cytogenetic/cytogenomic techniques such as karyotyping, fluorescence *in situ* hybridization (FISH) or chromosomal microarray analysis (CMA). However, these techniques are limited by the need for skilled personnel as well as significant time and labor, making them cost-prohibitive for some patients. Optical genome mapping (OGM) addresses these limitations and allows for the accurate identification of clinically significant SVs using a novel, high throughput, inexpensive methodology. In a single assay, OGM offers a significantly higher resolution than karyotyping with comprehensive genome-wide analysis comparable to CMA and the added unique ability to detect balanced SVs that are missed by microarray. Here, we report the performance of OGM in a cohort of 100 AML cases, which were previously characterized by karyotype alone or karyotype and FISH. CMA was performed as an additional test in some cases. OGM identified all the clinically relevant SVs and CNVs reported by these standard cytogenetic methods. Moreover, OGM identified clinically relevant SVs in 11% of cases that had been missed by the routine methods. In 24% of cases, OGM refined the underlying genomic structure reported by traditional cytogenomic testing (13%), identified additional clinically relevant variants (7%) or both (4%). Three of 48 (6.25%) cases reported with normal karyotypes were shown to have cryptic translocations involving gene fusions. Two of these cases included fusion between *NSD1-NUP98*. Based on the comprehensive genomic profiling of the AML patients in this multi-institutional study, we recommend that OGM be considered as a first-line test for detection and identification of clinically relevant SVs.

## Introduction

Acute myeloid leukemia (AML) is the most common acute leukemia in adults with approximate incidence of 3-5 cases per 100,000 individuals per year and is characterized by rapid abnormal proliferation and differentiation of a clonal population of myeloid stem cells^1,2^. All AML cases carry somatic sequencing variants, approximately half of which are large genomic rearrangements or structural variations (SVs) detectable by karyotyping, chromosomal microarrays (CMA) or fluorescence *in situ* hybridization (FISH)^3,4^.

Given the consistent correlation of clinical outcomes with specific mutation classes, the World Health Organization (WHO), National Comprehensive Cancer Network (NCCN) and European Leukemia Net (ELN) agencies developed recommendations for diagnosis and management of AML in adults based on the spectrum of somatic single nucleotide variants and SVs. While sequencing variants and small structural variants can be identified by targeted next generation sequencing (NGS), karyotyping and FISH are the standard of care technologies for the detection of well characterized SVs, including chromosomal translocations, inversions and copy number variants (CNVs) in AML^1,3,5^. However, classical karyotyping techniques have the limit of resolution of only 10 Mb and some SVs are cryptic and cannot be identified regardless of the limit of resolution. FISH has demonstrated higher sensitivity than karyotyping but has the limited utility of only evaluating the specific regions or targets of interest, thus requiring large FISH panels for a comprehensive evaluation of all clinically significant abnormalities in AML. CMA has proven to be useful for detecting CNVs that are beyond the resolution of karyotyping but is ineffective for the detection of balanced SV events such as inversions and translocations. These balanced SVs frequently generate gene-fusion products, knowledge of which often informs prognosis and guides potential therapeutic regimens^6,7^.

Given the individual limitations of karyotyping, FISH, and CMA, a single diagnostic assay that identifies all clinically significant structural variants is highly desirable. Different methods have been developed for the clinical diagnostics field in the past decade, including NGS^8^, whole-genome mate-pair sequencing^9^ (MPseq) and Anchored Multiplex PCR^10^ (AMP). However, NGS suffers from its inability to identify SVs associated with redundant elements or unmappable regions as well as the requirements of expensive instrumentation, complex bioinformatics pipeline and qualified healthcare personnel to interpret data generated from the clinical samples. AMP methods have been shown to have clinical utility but require *a priori* information and do not provide genome-wide analysis, thereby missing a number of genomic aberrations outside the scope of these methods.

Optical Genome Mapping (OGM) with Bionano Genome Imaging has demonstrable advantage in detection and identification all classes of SVs in the human genome^11–14^. OGM allows for high throughput, accurate and inexpensive testing of different cancer types for the identification of clinically relevant SVs within a single assay^15–18^. Specifically, OGM has greater sensitivity and resolution compared to karyotyping, can interrogate the whole genome in contrast to FISH and can detect balanced events that are missed by CMA^19,20^.

We evaluated the performance of OGM in a cohort of 100 bone marrow or peripheral blood specimens from patients with a diagnosis of AML. Clinical samples were initially received and processed at eight separate CLIA/CAP-certified laboratories using routine methods ordered by a referring clinician (karyotype, FISH and/or CMA). The results of this study demonstrated that Optical Genome Mapping not only detected all the structural variants identified by standard of care routine cytogenetic techniques but also provided superior accuracy and greater insight into the precise structure of the SVs compared to conventional chromosome studies. Timely detection of pathogenomonic findings in patients with AML is critical for accurate disease management and prognosis. With a fast turnaround time and elimination of the need of three different technologies to reach a laboratory diagnosis, OGM provides an attractive alternative to conventional technologies. Additionally, OGM was able to identify clinically significant SV calls in 11% of cases that were missed by conventional methodologies. Our study demonstrates that OGM has the potential to be the standard of care methodology for cytogenomic evaluation of patients with AML. Moreover, by identifying previously unrecognized SVs, OGM could play a significant role in the identification of targetable genomic aberrations for novel breakthrough therapeutic treatment options.

## METHODS

### Samples

Peripheral blood or bone marrow (BM) samples were obtained at diagnosis from AML patients who were subsequently treated using intensive chemotherapy (3+7). Patients were recruited under Institutional Review Board (IRB) protocols approved at each institution (Augusta University-HAC # 611298; Columbia University Irving Medical Center-IRB-AAAS0105; Fred Hutch Institutional Review Board-IR File#7067; Legacy Health IRB exempt; Mayo Clinic-17-003542; Pathgroup, Western IRB exempt; Penn State College of Medicine-#2000-186). Mononuclear cells (MNCs) were isolated by density gradient separation (Ficol-Paque, GE Healthcare Life Sciences, USA) and frozen for later use. All samples underwent karyotype analysis at a CLIA/CAP-certified clinical laboratory and in some cases, as indicated, FISH and/or CMA was performed.

### Optical Genome Mapping

Ultra-high molecular weight (UHMW) DNA was extracted from peripheral blood or bone marrow aspirates (BMA), largely following the manufacturer’s protocols (Bionano Genomics, USA). A minority of samples were cells separated from crude bone marrow by Ficoll separation, preserved at −80°C in freezing media and then washed with extra Cell and RBC Lysis Buffers pre-isolation. The WBC were digested with Proteinase K and RNAse A. DNA was precipitated with isopropanol and bound with nanobind magnetic disk. Bound UHMW DNA was resuspended in the elution buffer and quantified with Qubit dsDNA assay kits (ThermoFisher Scientific, USA).

DNA labeling was performed following manufacturer’s protocols (Bionano Genomics, USA). Standard Direct Label Enzyme 1 (DLE-1) reactions were carried out using 750 ng of purified high molecular weight DNA. Labeled DNA was loaded on Saphyr chips (Bionano Genomics, USA) for imaging. The fluorescently labeled DNA molecules were imaged sequentially across nanochannels on a Saphyr instrument. Effective genome coverage of approximately 300X was achieved for tested samples. All samples were evaluated based on molecule quality metrics. Specifically, the recommended values for reference genome GRCh38 molecule map rates were greater than 70% and molecule N50 (for molecules >150 kbp) values greater than 250kbp. In total, 78 samples met the recommended molecule quality metrics. Since these metrics were not established in a clinical setting, we assessed the remaining 22 samples using a hard map rate cutoff value of 55%.

Genome analysis was performed using software solutions provided by Bionano Genomics (Bionano Access and Bionano Solve). Rare Variant Analysis was performed to sensitively capture somatic SVs occurring at low allelic fractions. Briefly, molecules of a given sample dataset were first aligned against the public Genome Reference Consortium GRCh38 human assembly. SVs were identified based on discrepant alignment between sample molecules and GRCh38, with no assumption about ploidy. Consensus genome maps (*.cmaps) were then assembled from clustered sets of at least three molecules that identify the same variant. Finally, the genome maps were realigned to GRCh38, with SV data confirmed by consensus forming final SV calls. SVs generated by the Rare Variant Analysis were then annotated with known canonical gene set present in GRCh38 as well as estimated population frequency for each structural variant detected by comparing to a custom OGM database from Bionano Genomics.

Fractional copy number analysis was performed from alignment of molecules and labels against GRCh38 (alignmolvref). A sample’s raw label coverage was normalized against relative coverage from human controls, segmented, and baseline copy number (CN) state estimated from calculating mode of coverage of all labels. If chromosome Y molecules were present, baseline coverage in sex chromosomes was halved. With a baseline estimated, CN states of segmented genomic intervals were assessed for significant increase/decrease from the baseline. Corresponding gain and loss copy number variant calls were output. Certain SV and CN calls were masked if they occurred in GRC38 regions found to be high variance (gaps, segmental duplications, etc.).

### Assessment of Clinical Utility

The clinical utility of OGM was assessed in 2 primary ways. [1] Concordance with clinically significant SVs/CNVs reported by the CLIA/CAP laboratories using routine cytogenomic testing and [2] Identification of additional clinically significant SVs/CNVs not identified by routine testing. With the *a priori* knowledge that OGM reveals structural complexity undiscernible by karyotyping^21,22^, we sought to focus exclusively on SVs and CNVs of potential clinical significance. As such, all additional OGM findings were further filtered for overlap with AML specific genes (Supplementary Table S2) and AML specific FISH probe locations (Supplementary Table S2). Additional SV’s and CNVs identified were then further assessed using the following criteria: [1] variant absent in the Bionano Solve 3.5 database of human control samples, [2] size greater than 5 kb for insertions/deletions and greater than 5 Mb for CNVs.

### Curation of Additional Clinically Significant SVs/CNVs not Identified by Routine Testing

An expert review panel comprised of clinical molecular pathologists and clinical cytogeneticists (LB, RK, NS, MS) was formed to discuss the clinical significance of additional SVs identified by OGM after filtering all additional SVs and CNVs with AML specific genes and AML specific FISH probe locations (Supplementary Tables S2 and S3). The final list of additional SV’s and CNVs was compiled based on 2 tiers of clinical significance as follows: [1] Identification of one or more aberrations used in the ELN 2017 risk-stratification system or those that change the ELN risk status. [2] Identification of abnormalities that may or may not be specified in the ELN-2017 risk stratification system but for which sufficient published data demonstrates clinical significance ^23^.

### Concordance of Samples below 5% Allelic Fraction

With sufficient sample coverage, OGM has the potential to call SVs with allele fractions as low as 1% (internal unpublished data, Bionano Genomics). To emulate practical coverage that a clinical laboratory would likely attain in routine testing, we generated approximately 300x effective coverage for each sample which is predicted to identify SVs with allele fraction frequency of greater than 5%^24^. In routine cytogenetic analysis of 20 metaphase cells, heterozygous rare variants found in 1 or 2 cells (2.5%-5% allele frequency) are generally not considered to be clinically actionable. Therefore, SVs and CNVs that were identified by karyotype in ≤ 2/20 metaphases but not identified by OGM were not considered discordant (Supplementary Table 1).

### Data Analysis and Visualization

All SVs are displayed on a CIRCOS plot, showing abnormal fusions in the center, represented by lines connecting fusion positions, CNVs are shown in the inner ring (e.g. whole chromosome gains and losses), other SVs are shown in the next ring and chromosome specific cytobands are shown in the outer ring.

### Confirmation of Additional SVs and CNVs

Additional SVs and CNVs identified by OGM and deemed to be of potential clinical significance were confirmed by CMA and/or long-range PCR and/or Sanger sequencing as appropriate.

### OGM Nomenclature

Since OGM is a new technology, we modeled the OGM nomenclature on the microarray nomenclature described by ISCN 2016^25^. We have chosen ogm as the 3-letter prefix. Since microarray technology cannot detect balanced rearrangements, we have proposed a unique nomenclature using ISCN 2016 logic.

## RESULTS

We collected 100 adult acute myeloid leukemia samples that were previously sent for routine cytogenomic testing to a CLIA/CAP-approved clinical diagnostic laboratory. All 100 cases had G-banded karyotype results and a subset of these had FISH (19/100) and CMA (3/100) results (Supplementary Table 1).

To assist in calculating concordance of OGM findings with routine cytogenomic results, we stratified the cytogenomic results into five categories (Table 1): [1] Negative karyotype, [2] Aneuploidy and/or Aneusomy (partial aneuploidy or CNV), [3] Translocation and/or Inversion plus Aneuploidy/Aneusomy [4] Translocation/Inversion only, and [5] Complex with ≥ 3 abnormalities^26^. OGM identified all clinically relevant genomic abnormalities identified by routine cytogenomic analysis demonstrating 100% concordance for all 5 categories (Table 1). In addition, 3 karyotype negative cases were shown to have cryptic translocations involving gene-fusion partners. These included one case (Case ID 23, Table 2) with a t(3;12)(q26.2;12p13.2) translocation fusing *MECOM-ETV6* (Figure 4) and two cases (Case IDs 22 and 48, Table 3) with a t(5;11)(q35.3;p15.4) translocation fusing *NSD1-NUP98*. While the t(3;12)(q26.2;12p13.2) case was initially reported as 46,XX, subsequent FISH studies did identify a *MECOM* rearrangement, however, the translocation partner was never identified by routine cytogenomic testing. In 13% of cases, OGM either provided more accurate breakpoints in SVs or it resolved unknown cytogenetic elements reported by traditional cytogenetic testing (e.g. marker chromosomes or derivative chromosomes with material of unknown origin) by refining the underlying genomic structure. In 7% of cases, OGM identified additional clinically relevant SVs and/or CNVs (Table 3) and in 4% of cases OGM identified additional clinically relevant SVs and/or CNVs as well as resolved/refined the karyotype (Table 2). In total, OGM identified additional clinically significant SVs and/or CNVs in 11% of cases (Tables 2 and 3). Twenty-two samples did not pass the recommended molecule quality thresholds (discussed in methods). However, even with lower quality metrics, OGM identified all the clinically reported SVs and CNVs found by cytogenetics (Supplementary Table 1).

**Table 1.** Concordance rates achieved with Optical Genome Mapping for each type of Cytogenomic result reported per case by karyotype/FISH/CMA.

| METHOD | CYTOGENOMIC RESULT |  |  |  |  | Total |
| --- | --- | --- | --- | --- | --- | --- |
|  | Negative Karyotype | Structural Variants |  |  |  |  |
|  |  | Aneuploidy/ Aneusomy | Trans + Aneuploidy/ Aneusomy | Trans or Inv only | Complex > 3 SVs |  |
| Traditional Cytogenomics | 48 (-3*) | 16 | 2 | 25 | 9 | 100 |
| OGM | 45** | 16 (4)*** | 2 | 28** (2)*** | 9 (2)*** | 100 |
| OGM Concordance | 100% | 100% | 100% | 100% | 100% | 100% |

Trans: Translocation. Inv: Inversion. \* Number of negative karyotype cases confirmed to have additional SV findings. A negative number reduces the effective true number of cases in that category. \*\* The 3 additional SV findings in cases originally reported with a negative karyotype reduces the true negative karyotype case number down to 45 while concomitantly increasing the translocation/inversion case number up to 28. \*\*\* Number of cases within that category that were confirmed to have additional SV or aneuploidy/aneusomy findings.

**Table 2:** Refining Cytogenetic Breakpoints, Resolving Unknown Cytogenetic Elements, and Identifying Additional Clinically Relevant SVs and CNVs by OGM.

| Sample ID | Karyotype | FISH | OGM | Karyotype Predicted by OGM * | Overlapping Genes |
| --- | --- | --- | --- | --- | --- |
| 23 | 46,XY[20] | 3q+ (MECOM) (82%) | ogm[GRCh38]<br>**t(3q26.2;12p13.2)(169465166;11802643)** | 46,XY,t(3;12)(q26.2;p13.2) | MECOM-ETV6 |
| 51 | 46,XX,add(3)(p21),del(20)(p11.2q13.3)[1]* | monosomy 7 in 22%, MLL broken in 4% | ogm[GRCh38]<br>1p36.11p35.2(27043227_31075530)x1,<br>**3p22.1p14.3(41441584_57181743)x1,**<br>**7q21.3q36.1(96926735_152772221)x1,**<br>**20q11.22q13.13(33471428_51544362)x1,**<br>**21q22.11q22.12(32821007_36209023)x1** | 46,XX,del(1)(p36.11p35.2),<br>**del(3)(p14.3p22.1),**<br>**del(7)(q21.3q36.1),**<br>**del(20)(q11.22q13.13),**<br>**del(21)(q22.11q22.12)** | RUNX1 |
| 53 | 46,XX,del(6)(q21),add(17)(p13)[20] | ND | ogm[GRCh38]<br>**t(1p36.23;17p13.3)(8781440;2208744),**<br>**6q12q23.3(63771696_135674891)x1,**<br>**12p13.2(11508826_12722843)x1** | 46,XX,del(6)(q12q23.3),<br>**der(17)t(1;17)(p36.23;p13.3),**<br>**del(12)(p13.2)** | RERE-SMG6,ETV6 |
| 97 | 43-44,X,der(X)t(X;9)(q22;q22),add(2)(q31),add(3)(p21),del(3)(q11.1),add(4)(p14),add(5)(q11.2),-7,+8,-10,-11,add(12)(p11.2),add(14)(q22),-17,der(18)t(17;18)(q11.2;q23),add(19)(q13.3),add(20)(q11.2),+1,-3 mar[cp17]/84-89,idemx2[cp3] | ND | ogm[GRCh38]<br>t(1p22.3;Xq11.221)(86702476;131836283),<br>3p14.1p12.1(66723200_86246673)x1,<br>t(3q25.31;14q31.1)(155611536;80697114),<br>t(3q25.32;14q31.1)(157609007;79084975),<br>3p24.3p22.3(21748185_33387315)x1,<br>3q11.2q22.3(97753067_136337312),<br>t(4p15.33;12p11.2)(12582213;25901387),<br>4q24q27(104786659_122582963)x1,<br>5q15q33.3(94240748_160450226)x1,<br>**t(5q15;17p13.1)(94240748;7684753),**<br>(7)x1, (8)x3,<br>t(11q13.1;19q13.33)(65621402;48253924),<br>**16q21qter(63988035_90338345)x1,**<br>**18q12.1qter(28024374_80373285)x1,** 20q11.22qter(33120434_64444167)x1 | 45,XX,t(X;1),del(3)(p12.1p14.1),t(3;14)(q25.31;q31.1),del(3)(p22.3p24.3),del(3)(q11.2q22.2),t(4;12)(p15.33;p11.2),del(4)(q23q27),del(5)(q15q33.3),**t(5;17)(q15;p13.1),**-7,+8,t(11;19)(q13.1;q13.33),t(3;14)(q25.32;q31.1),**del(16)(q21q23.1),****del(18)(q12.1q23),**del(20)(q11.22q13.33) | TP53,CBFB,SETBP1 |

Bolded text indicates cytogenetic regions refined or resolved. Text in red indicates additional clinically relevant SVs and CNVs. \* The predicted karyotype does not include clonal information

**Table 3:** Identifying Clinically Relevant SVs and CNVs not Apparent by Routine Cytogenomic Testing by OGM.

| Sample ID | Karyotype Results | FISH | OGM Finding | OGM Predicted Karyotype | Overlapping Genes |
| --- | --- | --- | --- | --- | --- |
| 22 | 46,XY[20] | ND | ogm[GRCh38]<br>**t(5q35.3;11p15.4)(177215724;3743680)** | 46,XY,**t(5;11)(q35.3;p15.4)** | *NSD1-NUP98* |
| 48 | 46,XX[20] | ND | ogm[GRCh38]<br>**t(5q35.3;11p15.4)(177225609;3743680)** | 46,XX,**t(5;11)(q35.3;p15.4)** | *NSD1-NUP98* |
| 61 | 47,XX,+9[15]/46,XX[5] | ND | ogm[GRCh38]<br>(9)x3,<br>**inv(16p13.11q22.1)(15727754_67101201)** | 47,XX,+9,**inv(16)(p13.11q22.1)** | *NDE1/MYH11-CBFB* |
| 62 | 49,XY,+8,+9,+14[20] | ND | ogm[GRCh38]<br>(8)x3, **t(9p21.3;11q23.3)(22078859;118493942)**, (9)x3,<br>11q23.3qter(118606985_135086622)x3,<br>(14)x3 | 49,XY,+8,**t(9;11)(p21.3;q23.3)**,+9,+14 | *MLLT3-KMT2A* |
| 68 | 46,XY,t(3;5)(q25;q35)[20] | ND | ogm[GRCh38]<br>t(3q25.32;5q35.1)(158574030;171400357),<br>**21q21.1q21.3(32402419_36892141)x1** | 46,XY,t(3;5)(q25.32;q35.1),<br>**del(21)(q22.11q22.12)** | *NPM1-MLF1,RUNX1* |
| 75 | 46,XX,t(8;21)(q22;q22)[20] | ND | ogm[GRCh38]<br>**t(4q26;5q21.3)(116664779;109795032)**,<br>4q26q35.2(116717489_187080.531)x3,<br>5q22.1q35.3(109795032_180908224)x1,<br>**8q21.3q22.1(92067075_94847624)x1**,<br>t(8q21.3;21q22.12)(92068759;34855785) | 46,XX,**der(5)t(4;5)(q26;q21.3)**,<br>t(8;21)(q21.3;q22.12),<br>**del(8)(q21.3q22.1)** | *RUNX1T1-RUNX1,EGR1* |
| 100 | 47,XX,der(3)add(3)(q27)ins(3)(3;?)(p21;?),del(5)(q13q35),dic(7;12)(q11.2;p11.2),+9,add(10)(q11.2),del(17)(q24),+22[13]/47,idem,add(2)(q37),ad d(4)(q21)[3]/44,XX,der(1)t(1;6)(q11.2;p11.2),add(2)(q24),der(3)add(3)(q27)ins(3)(3;?)(p21;?),del(5)(q13q35),dic(7;12)(q11.2;p11.2),add(10)(q11.2),del(17)(q24)[4] | loss of 5q, 7q, and AML | ogm[GRCh38]<br>t(2q37.1;4q22.1)(233962907;92654205),<br>t(3p21.31;10q11.22)(57139125;48365212),<br>5q14.3q35.1(85426887_169497669)x1,<br>7p14.1p11.2(39952265_57740993)x1,<br>7q11.21qter(65027424_159345973)x1,<br>(9)x3, **17p13.1(7642059_7693432)x3**,17q21.31qter(45633925_83257441)x1, (22)x3 | 48,XX,t(2;4)(q37.1q22.1),t(3;10)(p21.31;q11.22),del(5)(q14.3q35.1),del(7)(p11.2p14.1;q11.21q36.3),+9,**dup(17)(p13.1)**,del(17)(q21.31q25.3),+22 | *TRPM8;GRID2,TGM4;FAM149B1,TP53* |

Text in red indicates additional clinically relevant SVs and CNVs. \* The predicted karyotype does not include clonal information

### Detection of Aneuploidy/Aneusomy and SVs – Concordant Samples

OGM detected all 16 cases reported by routine cytogenomic testing with aneuploidy and/or aneusomy (Supplementary Table 1). Large copy number changes are easily visualized in the circus plot output or the whole genome CNV profiles (Figure 1). Similarly, all 36 cases with translocations and/or inversions identified by G-banded karyotype were detected by OGM (Supplementary Table 1). Classic AML translocations, such as the t(8;21)(q22;q22) translocation fusing *RUNX1T1-RUNX1*, rely on both karyotype and FISH for definitive identification and these were identified in a single step by OGM. Figure 2 shows an example (case 933) where G-banded karyotyping revealed two distinct translocations t(8;21)(q22;q22) and t(10;13)(q22;q12) (Figure 2A). The two translocations are easily observed on the circus plot (Figure 2B) and the *RUNX1T1-RUNX1* fusion is revealed by assessment of the linear genome browser view, showing mapping to the junction regions (Figure 2C). The t(10;13)(q22;q12) translocation does not overlap any known genes and thus this rearrangement does not yield gene to gene fusion. In these specific cases, FISH was not performed and the *RUNX1T1-RUNX1* fusion was assumed based on the karyotype. While this assumption is accurate in most instances, additional complexities such as a partial deletion of *RUNX1T1* accompanying the translocation/fusion, as seen in case 75 (Table 3), would not be identified and could potentially yield a false negative if only FISH was performed on interphase nuclei. Other examples of recurrent AML specific translocations involving chromosomes 3;5 (*NPM1-MLF1*) and 9;11 (*KMT2A-MLLT3*) resulting in gene fusions are shown in Supplementary Figure 3.

**Figure 1:**
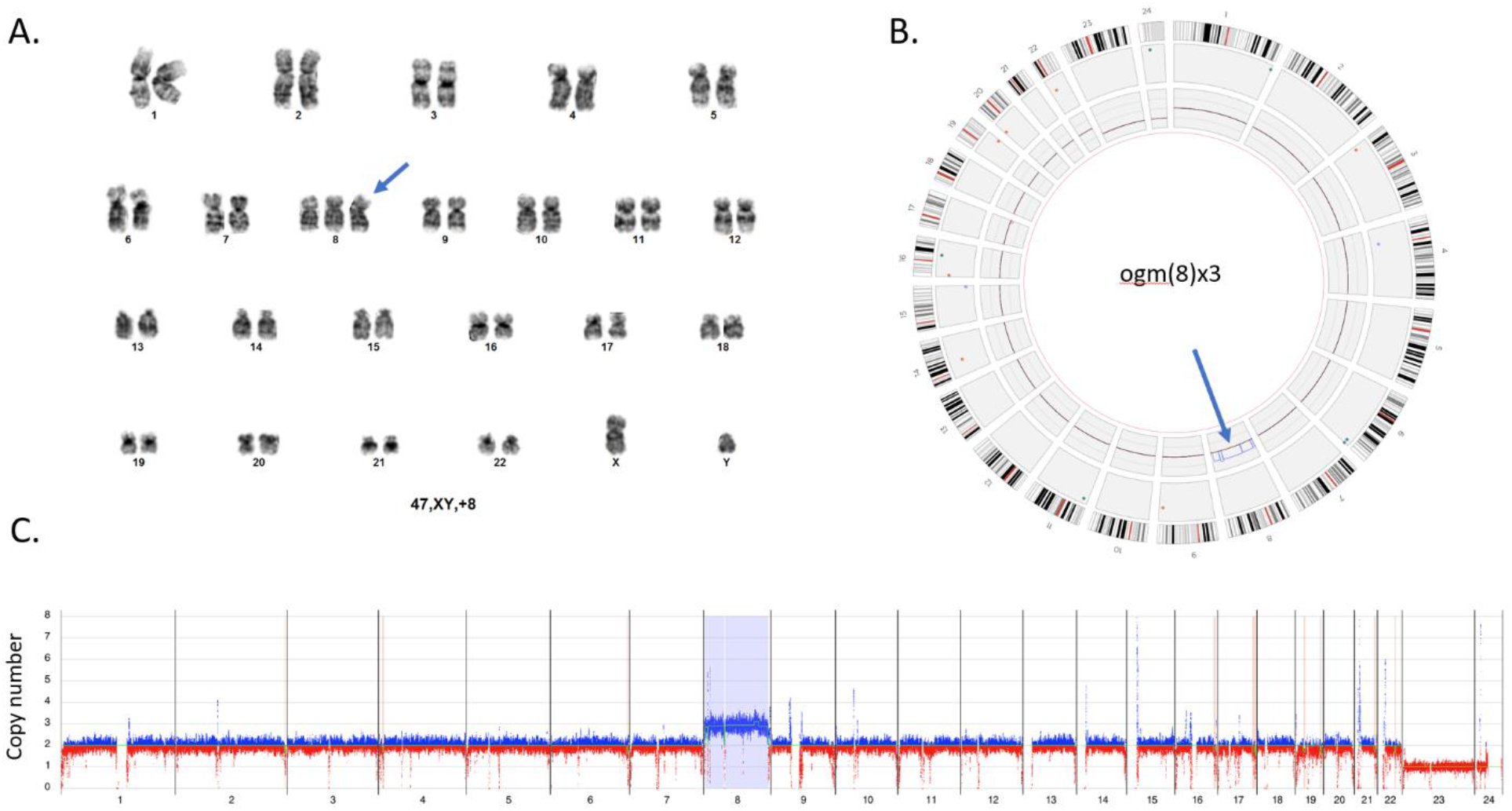
Identification of Aneuploidies by OGM. **A**. Standard G-banded karyotype with trisomy 8. **B**. Visualization of trisomy 8 in the circos plot output. The extra copy of chromosome 8 is shown within the inner layer of the circos plot (blue arrow). **C**. Whole genome CNV profiles generated with Bionano Access indicates a gain of chromosome 8. Y axis shows copy number range from 0-8 for each chromosome. X axis shows molecules with an increased copy number in blue and those with a decreased copy number in red. The light blue region highlighted for chromosome 8 indicates a significant difference from the baseline thus flagging a gain of chromosome 8. All chromosomes except the sex chromosomes (X,Y)x1 and chromosome (8)x3 are present in 2 copies. Sample – 16. Note: chromosome X=23, and Y=24.

**Figure 2:**
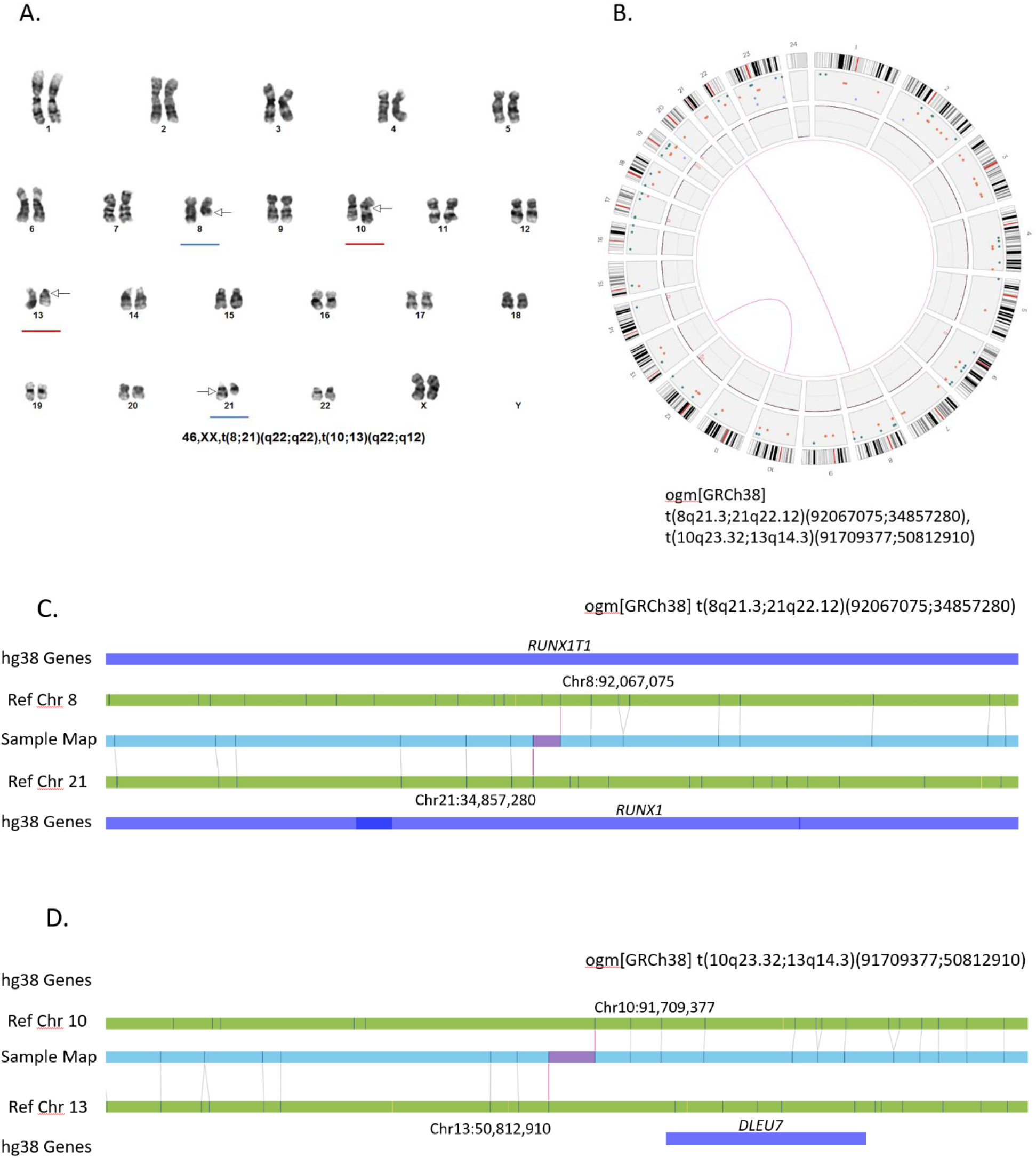
Identification of Balanced Translocations by OGM. **A**. Standard G-banded karyotype showing t(8;21)(q22;q22) (blue underline) and t(10;13)(q22;q12) (red underline). **B**. Circos plot demonstrating a translocation between chromosomes 8 and 21 (pink line connection between the two chromosomes) and a second translocation between chromosomes 10 and 14 (pink line connection between the two chromosomes). The Bionano Access software allows fine zooming on individual chromosomes to attain finer detail of the cytogenetic breakpoints involved. **C**,**D**. Linear genome browser representation of the two identified translocations with Bionano Access. GRCh38 reference chromosomes with OGM label patterns are shown in green. Assembled patient sample maps with label patters are shown in light blue. Label alignments between two maps are shown in grey strings. Translocation breakpoints are highlighted in purple. Overlapping genes are shown in blue. Sample – 77.

Inversion events may also affect critical genes or yield gene fusions and Figure 3 shows two examples of classic AML inversions which can be detected by routine cytogenetic analysis. OGM detected 2 cases with the classic inv(3)(q21.3q26.2) inversions, associated with poor outcome and placing the inverted segments adjacent to *MECOM* on chromosome 3 (example shown in Figure 3A). Five cases with the classic inv(16)(p13.11q22.1) fusing *MYH11-CBFB* were reported by routine testing and detected by OGM (example shown in Figure 3B). Both inversions are called based on two map alignments within each chromosome. For example, sample maps (blue) in Figure 3 show both linear and inverted alignments around the inversion breakpoints, where half aligns to one location of the reference genome and the second half aligns to another part of the reference (same chromosome), but in inverted orientation. The resultant gene fusion resolution obtained with OGM is orders of magnitude higher than attainable by karyotyping.

**Figure 3:**
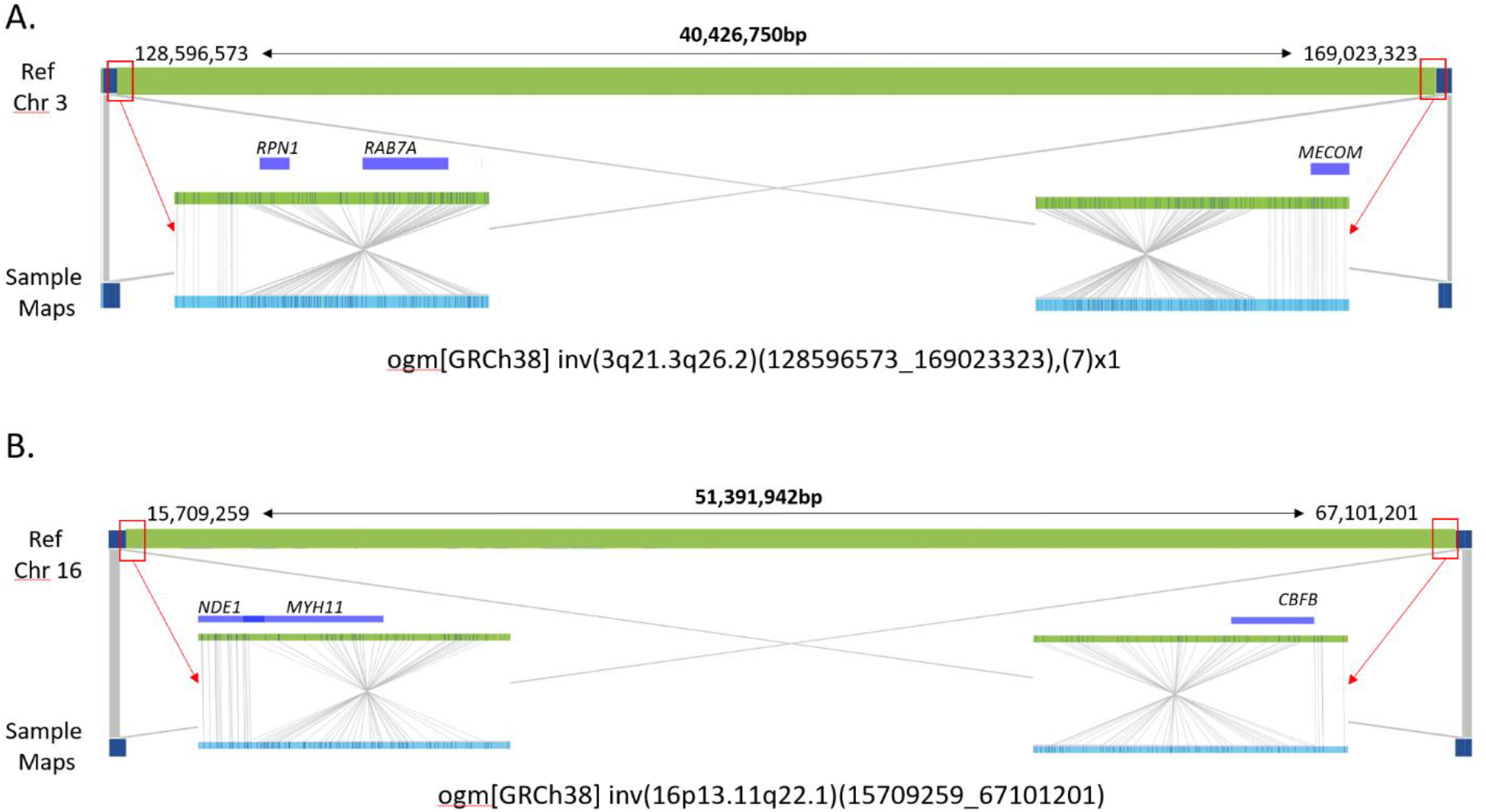
Identification of Balanced Inversions by OGM. **A**. Large chromosome 3 inversion identified in AML samples – 82,83. **B**. Large chromosome 16 inversion identified in AML samples – 84-89. GRCh38 reference chromosomes with OM label patterns are shown in green. Assembled sample maps with label patters are shown in light blue. Label alignments between two maps are shown in grey strings. Overlapping genes are shown in blue.

**Figure 4:**
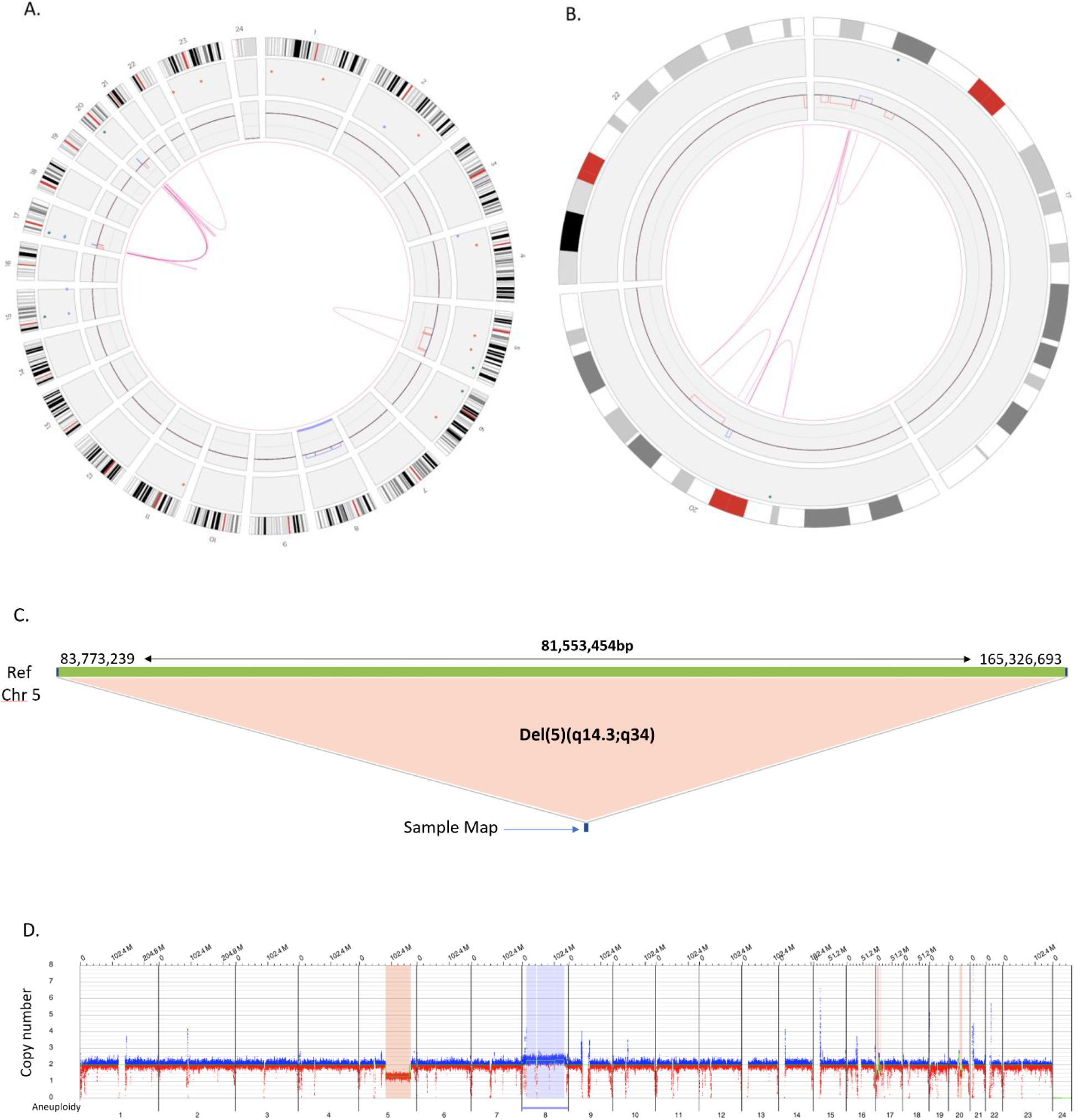
Production of an Accurate Cytogenomic Karyotype by Refining Cytogenetic Breakpoints and Resolving Unknown Cytogenetic Elements by OGM. Routine karyotyping in sample 93 was reported as: 46,XX,del(5)(q13q33)[4]/45,idem,add(17)(p11.2),-20, add(22)(q13)[6]/ 46,idem,+8[10]. OGM refined the breakpoints of the 5q deletion to del(5)(q14.3q34) and identified the additional material on chromosomes 17 and 22 to derive from different regions of chromosome 20. The karyotype indicated that chromosome 20 is absent from the cell, however, OGM indicates that most of chromosome 20 is indeed present but fractured and reassembled into different regions of chromosomes 17 and 22. **A**. Circos plot showing a whole genome view of the multiple genomic rearrangements (pink lines) and copy number profiles (inner circle blue boxes indicate gains and red boxes indicate deletions). **B**. Circos plot showing a selected chromosome view of the complex genomic rearrangements (pink lines) and copy number profiles (inner circle blue boxes indicate gains and red boxes indicate deletions) between chromosomes 17, 20 and 22. **C**. Fine mapping of the chromosome 5 deletion indicates a large 81.5Mb deletion on the q arm of chromosome 5 between genomic coordinates 83,773,239 and 165,326,693 (human genome build GRCh38). **D**. Whole genome CNV profile showing interstitial deletion on 5q as well a gain of chromosome 8.

### Assembling the Complete Karyotype by Refining Cytogenetic Breakpoints and Resolving Unknown Cytogenetic Elements

In 13 cases, OGM either provided more accurate breakpoints in SVs or it resolved unknown cytogenetic elements reported by traditional cytogenetic testing such as marker chromosomes or chromosomes with additional material of unknown origin (Table 4). The majority of these cases involved identifying the origins and structural nature of the additional material of unknown origin. Many of these turned out to be cryptic translocations. For example, in case 49 (Table 4), additional material of unknown origin was observed by karyotype to be present at band 8p23. OGM revealed that the additional material derived from chromosome 12 resulting in an unbalanced translocation, predicted by OGM to be der(8)t(8;12)(p23.2;q23.1) (Table 4). Figure 4 illustrates a complex karyotype with at least 5 abnormalities (case 93, Table 4). The CIRCOS plot generated by Bionano access revealed the genomic complexity of the case. These included an interstitial deletion of chromosome 5q, a complete gain of chromosome 8, and numerous rearrangements between chromosomes 17, 20, and 22 (Figure 4A). Detailed assessment of the rearrangements involving chromosomes 17, 20 and 22 revealed the underlying genomic architecture of the structural rearrangements (Figure 4B).

**Table 4:** Refining Cytogenetic Breakpoints and Resolving Unknown Cytogenetic Elements by OGM.

| Sample ID | Karyotype | FISH | OGM | Karyotype Predicted by OGM * | Overlapping Genes |
| --- | --- | --- | --- | --- | --- |
| 47 | 46,XX[20] | partial deletion of TET2 (82.0%) | ogm[GRCh38] 4q24(104950756_105321326)x1 | 46,XX,**del(4)(q24q24)** | *TET2* |
| 49 | 44,XY,del(1)(q42),-7,add(8)(p23),-12[18]/44,idem,t(4;5)(p14;q13)[2] | ND | ogm[GRCh38] 1q41(217038801_217902115)x1, 1q41(219849117_220554040)x1,(7)x1,<br>**t(8p23.2;12q23.1)(2741116;96775720)**,<br>12p13.2p12.3(10049506_15008329)x1,<br>12p12.3q11(19135976_34717946)x1,<br>12q21.31(82305245_84374961)x1,<br>12q21.31q21.32(86295391_87098300)x1,<br>12q21.33q23.1(89358577_96763295)x1,<br>12q24.13q24.31(113619222_120327577)x1 | 44,XY,del(1)(q41),-7,<br>**der(8)t(8;12)(p23.2;q23.1)**, -12 | multiple genes (chr1, chr7, chr12) |
| 50 | 46,XX,add(2)(q33)<br>[8]/46,XX,del(5)<br>(q22q33)[5]/45,X,-<br>X,t(9;22)(q34;q11.2)[2]/46,XX[5] | ND | ogm[GRCh38]<br>t(2q33.3;q28.1)(206925289;124391375),<br>2q33.3qter(206642387_242193529)x1,<br>4q28.1qter(124443604_190214555)x3,<br>t(5p14.1;q23.2)(28887896;126172032) | 46,XX,der(2)t(2;4)(q33.3;q28.1),<br>del(2)(q33.3;q37.3),<br>t(5;5)(p14.1;q23.2) | multiple genes (chr2, chr4) |
| 52 | 46,XY,add(5)(q31)<br>[11]/46,XY[9] | ND | ogm[GRCh38]<br>t(5q23.1;q35.3)(119366177;87045338),<br>5q23.1qter(119366177_181538259)x1,<br>12q35.3qter(87045338_133275309)x3 | 46,XY,del(5)(q23.1;q35.3),<br>t(5;12)(q23.1;q21.32),<br>dup(12)(q21.32;q24.33) | EGR1,NPM1,DDX41,FLT4,TNFAIP<br>8,PTPN11 |
| 54 | 46,XX,add(7)(q11.2)[19]/47,idem,<br>+8[1] | ND | ogm[GRCh38]<br>7p11.2p21.3(8948293_57803971)x3,<br>7q11.2qter(63727644_159345973)x1 | 46,XY,dup(7)(p11.2p21.3),<br>del(7)(q11.2q36.3) | multiple genes (chr7) |
| 56 | 44,XY,add(9)(p13),<br>-16,-17,+mar[cp3] | ND | ogm[GRCh38] t(8p12;q23)(35986181;11749433),<br>t(9p23;q11.2)(11417993;13925135),<br>16p13.3(887824_6732043)x1,<br>16p13.13p11.2(11034760_32222510)x3,<br>16q11.2qter(38277017_90338345)x1,(Y)x0 | 45,X,-Y,t(8;9)(p12;p23),<br>der(9)t(9;21)(p23;q11.2),<br>dup(16)(p11.2p13.13),<br>del(16)(q11.2q24.3) | CBFB |
| 58 | 48,XY,+2,add(2)<br>(p11.2),add(3),<br>+14[20] | ND | ogm[GRCh38]<br>2p21qter(42997509_242193529)x3,<br>t(2p21;q26.2)(42967231;169765605),<br>3q26.2qter(169795957_198295559)x3,<br>(14)x3 | 48,XY,der(2)t(2;3)(p21;q26.2),+2,+14 | ACTR3 |
| 89 | 47,XY,t(16;16)(p13.1;q22),+22[20] | ND | ogm[GRCh38]<br>inv(16p13.1q22.1)(15727754_67101201), (22)x3 | 47,XY,inv(16)(p13.1q22.1),+22 | NDE1/MYH11-CBFB |
| 90 | 46,XX,add(7)(q22),<br>inv(16)(p13.1q22)<br>[19]/46,XX[1] | ND | ogm[GRCh38]<br>7q22.1q36.1(101635788_152284980)x1,<br>inv(16p13.1q22.1) (15727754_67101201) | 46,XX,del(7)(q22.1q36.1),<br>inv(16)(p13.1q22.1) | NDE1/MYH11-CBFB |
| 93 | 46,XX,del(5)(q13q33)[4]/45,idem,<br>add(17)(p11.2),-<br>20,add(22)(q13)[6]/46,idem,+8[1<br>0] | deletion of<br>EGR1 (60%),<br>TP53<br>(12.5%) and<br>trisomy<br>8(25%) | ogm[GRCh38]<br>5q14.3q34(83773239_165326693)x1,(8)x3,<br>t(17p13.1;q20q11.21)(7746369;45022656),<br>20q11.22q12(34253897_41607985)x1,<br>t(20q13.12;q23.33)(45037946;49612939) | 47,XX,del(5)(q14.3q34),+8,<br>der(17)t(17;20)(p13.1;q11.21),<br>del(20)(q11.22;q12)<br>der(20),t(20;22)(q13.12;q13.33) | EGR1;CHD3-HM13,TP53,STK4,TP53 |
| 94 | 41,XX,-<br>5,add(6)(q13),der(7;16)(p10;q10),<br>-11,-16,-17,add(17)(p11.2),-<br>18,add(21)(p11.2),<br>add(22)(p11.2)[18]/46,XX[2] | ND | ogm[GRCh38]<br>t(5q15;q17p11)(94299163;22306474), 5q11.2,q13q<br>14,q31,q333,q35)x1,6q16qter(96899849_170805<br>979)x1,7q11q31.33(62308387_126792419)x1,<br>7q33qter(136139026_159345973)x1, 11q14q22q<br>23q24x3, (16)x1, 17p11p13q11q21.31x1, (18)x1,<br>(22)x3 | 44,XX,t(5;17)(q15;p11),del(5)(q11.2,<br>q13q14,q31,q333,q35),del(6)(q16qter),<br>del(7)(q11qter),del(11)(q22q23),<br>dup(11)(q14,q22,q23,q24),<br>-16,del(17)(pterp13,q11q21.31)<br>-18,dup(22)(q11qter) | KIAA0825, multiple genes(chr5,<br>chr6, chr7, chr11, chr17, chr18,<br>chr22) |
| 95 | 52-56,XX,<br>add(3)(p21),add(5)(q11.1),add(8)(<br>q24.1),der(17)add(17)(p11.2)<br>add(17)(q25),-21,-22,+9-<br>16mar[cp7]/52-57,sl,-<br>11,add(13)(p11.2)[cp11]/46,XX[2] | ND | ogm[GRCh38]<br>t(3p24.1;q23.2)(34194457;127066149),<br>5q11.2q31.3(57794661_142239385)x1,<br>t(5q11.2;q21.2)(52646574;25433106),<br>t(5p13;q11q23)(36640261;116213934),<br>t(5q33;q25.3)(156834899;81874518),<br>t(13q11;q21.31)(18243591;26439456),<br>17pterp13.3(0_1347447)x1 | 46,XX,der(3)t(3;5)(p24p26;q23.2),<br>del(5)(q11.2q31.3),der(5)t(5;21)(q11.2q<br>21.2),t(5;11)(p13;q23),t(5;17)(q33;p11.2<br>),t(5;17)(q33;q25.3),t(13;21)(q11;q21.3),<br>del(17)(p13.1p13.3) | KMT2A,MYH10,TP53 |
| 96 | 45~46,XY,-<br>5,der(5)add(5)(p13)add(5)(q22),<br>add(9)(q22),add(10)(q22),-<br>12,del(17)(p11.2),der(17)t(11;17)(<br>q13;p11.2),add(22)(q13),+0~1mar<br>[cp9] | 11q+<br>(17.0~21.5<br>) and 17p-<br>(96.0%) | ogm[GRCh38]<br>t(5q31.1;q22.31)(134127916;93564274),t(5p13.<br>2;q22q12.2)(37001293;29364105),t(10p12.31;23p<br>11.4)(21549464;41366811),17pterp13.3(0_17415<br>988)x1 | 46,XY,t(5;9)(q31.1;q22.31),<br>der(5)t(5;22)(p13.2;q12.2),<br>t(10;23)(p12.31;p11.4),<br>del(17)(p11.2p13.3) | TCF7,NIPBL,AP1B1,DDX3X,MLLT<br>10;DDX3X,TP53 |

**Bolded text indicates cytogenetic regions refined or resolved. \* The predicted karyotype does not include clonal information**

### Identification of Additional Clinically Relevant SVs and CNVs

Our expert panel identified 11 cases with additional clinically significant SVs and/or CNVs that were not detected by routine testing methods (Tables 2, 3). All OGM findings identified in these 11 cases were confirmed by PCR and/or CMA (Supplementary Table S1). In 4/11 of these cases, OGM refined the existing karyotype as well as identified additional clinically significant genomic variants (Table 2). In 7/11 of these cases, OGM revealed completely new clinically significant aberrations that were not identified by routine testing (Table 3). The SVs included cryptic translocations such as t(5;11)(q35.3;p15.4) [cases 22 and 48, Table 3], t(3;12)(q26.2;p13.2) [case 23, Table 2) and t(9;11)(p21.3;q23.3) [case 62, Table 3). CNV changes were also identified (e.g. ogm[GRCh38] 16q21qter(63988035_90338345)x1, 18q12.1qter(28024374_80373285)x1 [case 97, Table 2), and, ogm[GRCh38] 21q21.1q21.3(32402419_36892141)x1 [case 51, Table 2]). In sample 75 (Table 3), a cryptic unbalanced der(5)t(4;5)(q26;q21.3) was uncovered by OGM in addition to the classic t(8;21)(q22;q22) translocation fusing *RUNX1T1-RUNX1* (Figure 5). This finding was particularly relevant as the loss of 5q could possibly lead to changes in patient management and prognosis. In addition, part of *RUNX1T1* is also deleted thus potentially creating a novel *RUNX1T1-RUNX1* fusion. The significance of this is uncertain but fusion predicted by routine testing would not be accurate.

**Figure 5:**
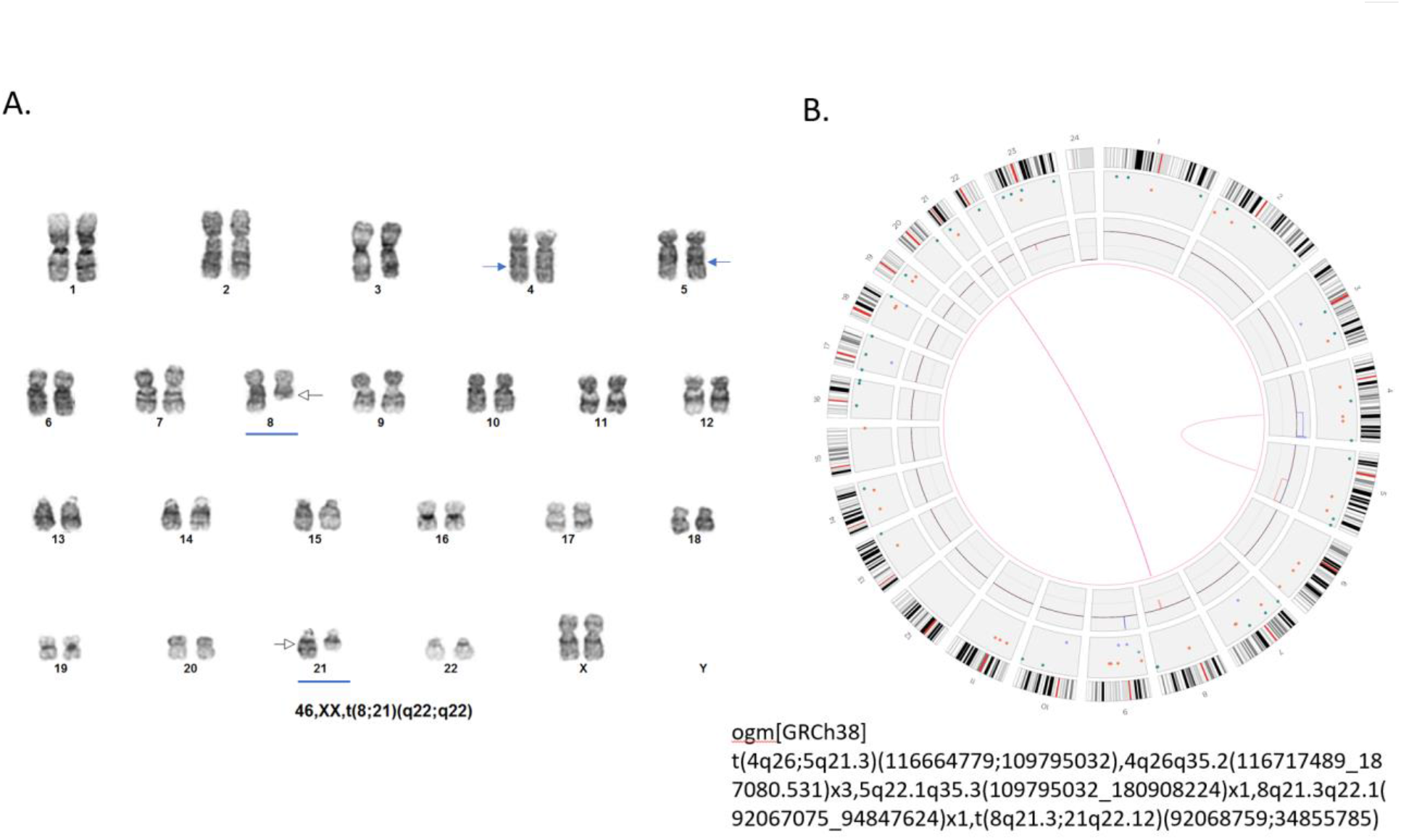
Identification of a Clinically Significant Large but Cryptic Unbalanced Translocation by OGM. **A**. Routine GTG karyotyping in sample 75 was reported with a single translocation as: 46,XX,t(8;21)(q22;q22) (Blue arrows) **B**. Circos plot showing the unbalanced t(8;21) (pink lines) with a cryptic deletion involving the *RUNX1T1* gene. Circos plot also reveals a second translocation between the long arms of chromosomes 4 and 5 (pink lines). The t(4;5) appears unbalanced as indicated by the copy number profiles where the inner circle blue box indicate a gain of distal 4q and the red box indicate a loss of distal 5q. deletions). Fine mapping places the 4q breakpoint at q26 and shows that the gain is approximately 70.4 Mb in size. The 5q breakpoint is mapped to band q22.1 and has an approximate size of 71.1 Mb. Therefore, although the unbalanced regions are extremely large, they represent a swap of similar appearing banded material of virtually identical size and would therefore be considered a cryptic rearrangement.

### Identification of Novel SVs and CNVs

Multiple novel uncurated SVs and CNVs were identified in this study. These are all listed in Supplementary Table S3 and may serve as potential candidates for future research.

## Discussion

This work demonstrates the performance of OGM for the analysis of a sizable number of clinical AML samples that contain in total a wide representation of currently known clinically relevant chromosomal abnormalities occurring in a cell fraction of at least 10%. We have shown that optical genome mapping matches the diagnostic scope achievable by routine cytogenomic methods and adds significant new information in upwards of 11% of cases. Furthermore, the performance of OGM surpasses even the combination of multiple tests and presents a much more refined and simplified workflow with additional cost benefits. Indeed, in 24% of cases, OGM assembled a more complete and accurate karyotype by refining cytogenetic breakpoints, resolving unknown cytogenetic elements, and detecting additional clinically significant SVs and CNVs.

Karyotyping provides a whole genome analysis of single cells and has been the standard of care for AML patients for decades. This study demonstrated several advantages of OGM compared to karyotyping with no obvious deficiencies in performance. Both methods effectively detect balanced and unbalanced chromosomal abnormalities such as duplications, deletions, aneuploidies, translocations, and inversions. While karyotyping is a single cell methodology and can provide information about clonality and subclone populations, OGM is a single molecule approach that can infer clonality based on allele fractions of SVs and CNVs. Since OGM utilizes single molecules derived from many cells, identification of SVs in a fraction of cells is more sensitive than routine analysis of 20 G-banded metaphases. This provides better statistical estimation of the clonal composition of the input material as previously demonstrated^22^. Moreover, since OGM directly interrogates the tumor without requiring cell culturing, as with karyotyping, the actual subclone frequency within the tumor can be calculated.

A distinct advantage of using OGM is the precise assignment of SV and CNV breakpoints indicating gene fusions, uncovering cryptic translocations and identifying CNVs below the resolution of standard G-banded karyotyping (<10 Mb). While these smaller CNVs can be detected by CMA or targeted FISH, CMA is not yet universally performed on AML specimens and only a limited number of FISH probes targeting specific AML gene fusions and/or hallmark abnormalities are typically performed. In the current study, all CNV’s <10Mb reported by FISH or CMA were detected by OGM (e.g. 370 Kb deletion involving *TET2*, case 47, Table 4). It is important to note that there is great variation in the FISH panels offered by clinical laboratories and the choice of panel often follows a laboratory or clinician specific algorithm. Each additional FISH panel adds extra healthcare costs to the patient. Break-apart probes are commonly used to identify abnormal gene-fusions. In many cases, the gene-fusion partners are identified by FISH but frequently the gene-fusion partner remains unidentified^27^. This study revealed that OGM reliably identifies gene-fusion partners, known and unknown in an unbiased manner. Examples include: *NPM1-MLF1, RUNX1-RUNX1T1, MECOM-ETV6*, and NSD1-NUP98 (Figure 3C, Supplementary Figure 3, Supplementary Table S1).

Recent NGS based approaches have been proposed and tested for replacement of traditional cytogenetics methods^28^. All NGS short read methods are challenged when dealing with highly repetitive regions that comprise a large fraction of the sequence reads. While whole genome sequencing detects most single nucleotide polymorphisms^29^, CNVs are also detected but generally with low resolution for size and position^30^. Furthermore, WGS cannot effectively detect many critical fusions and often cannot differentiate between translocations and insertions^31^. Since OGM leverages very long DNA molecules and scans long DNA molecules from the entire genome, the specificity of translocation detection is extremely high. OGM can detect all abnormal fusions except those with breakpoints in centromeres and other very long repeat structures such as the short arms of acrocentric chromosomes (no clinically actionable genes are located in the acrocentric short arms)^20^.

Adaptations of NGS have been applied to clinical cancer genomes especially in the form of panels. Anchored multiplex PCR (AMP) is one such method, which can detect point mutations as well as some fusions. This approach depends on the knowledge of one fusion partner with high precision and then can identify other fusion partners^10^. The approach is still limited to known targets and short sequence reads. Mate pair sequencing is a powerful technique for detection of genomic fusions in a whole genome approach^32^. Since paired sequence reads come from multiple sites adjacent to a fusion, some of the ambiguity of short read can be overcome. However, all NGS based approaches suffer from several critical limitations. Equipment price is a key consideration with the most cost-effective high throughput sequencers costing around $1 million and smaller less expensive sequencers ($100,000-$500,000) having higher per sample costs. Another common issue is the complexity of the sequence library preparation which requires many individual steps and often takes multiple days to conduct. Finally, data analysis complexity ranges from trivial in some targeted assays to incredibly complex and computationally expensive in other assays.

OGM requires only a short turnaround time, approximately 9 hours for DNA isolation and library preparation, with only 2 hours of hands-on time, and 24 hours for automated data collection. The assay can be conducted without significant specialized training by a laboratory technician that has experience with general molecular biology techniques.

There are some drawbacks to the OGM technique. Firstly, the OGM assay requires high molecular weight DNA isolated by specialized kits which precludes its use on most archival DNA banks and formalin-fixed-paraffin-embedded tissue. OGM is not a sequencing-based assay as it does not determine the sequence of individual base pairs and can therefore not identify single nucleotide variants. Finally, the throughput of the current OGM equipment is relatively low which makes implementation in a high-volume laboratory challenging. However, since the beginning of the current study, throughput has quadrupled and is expected to be even greater in the near future. On the other hand, none of the available NGS based methods can detect all the cytogenetic anomalies required for assessment of AML patients. Considering that abnormalities generally needing three independent assays for detection can be detected at once with OGM, the current OGM workflow represents a significant time and cost savings for potential clinical AML testing. Additionally, since OGM analytics are automated, standardized care across different testing laboratories can be achieved.

## Conclusion

We have demonstrated that optical genome mapping is equivalent in diagnostic scope to routine cytogenomic methods and adds significant new information in upwards of 11% of cases, including a potential increased diagnostic yield of 6.25% in cases reported with normal karyotypes (3/48). The results of this study indicate that OGM provides AML patients with a more comprehensive genomic analysis that can potentially influence clinical management. As such, OGM should be considered as a preferential replacement of current cytogenomic methodologies for detecting large SVs and CNVs. OGM has the potential to significantly advance our understanding of genotype/phenotype correlations by uncovering a more complete genomic karyotype which may ultimately lead to the development of alternate treatment options. Indeed, the novel findings uncovered by OGM will promote future research investigating their impact on patient outcome and possible therapeutic targets. Adoption of OGM into clinical laboratories should improve the diagnostic yields not only in AML patients but also in a much wider range of patients with other cancers.

## Data Availability

All the relevant data is available in the manuscript and supplementary files.

---

**Supplementary Figure 1:**
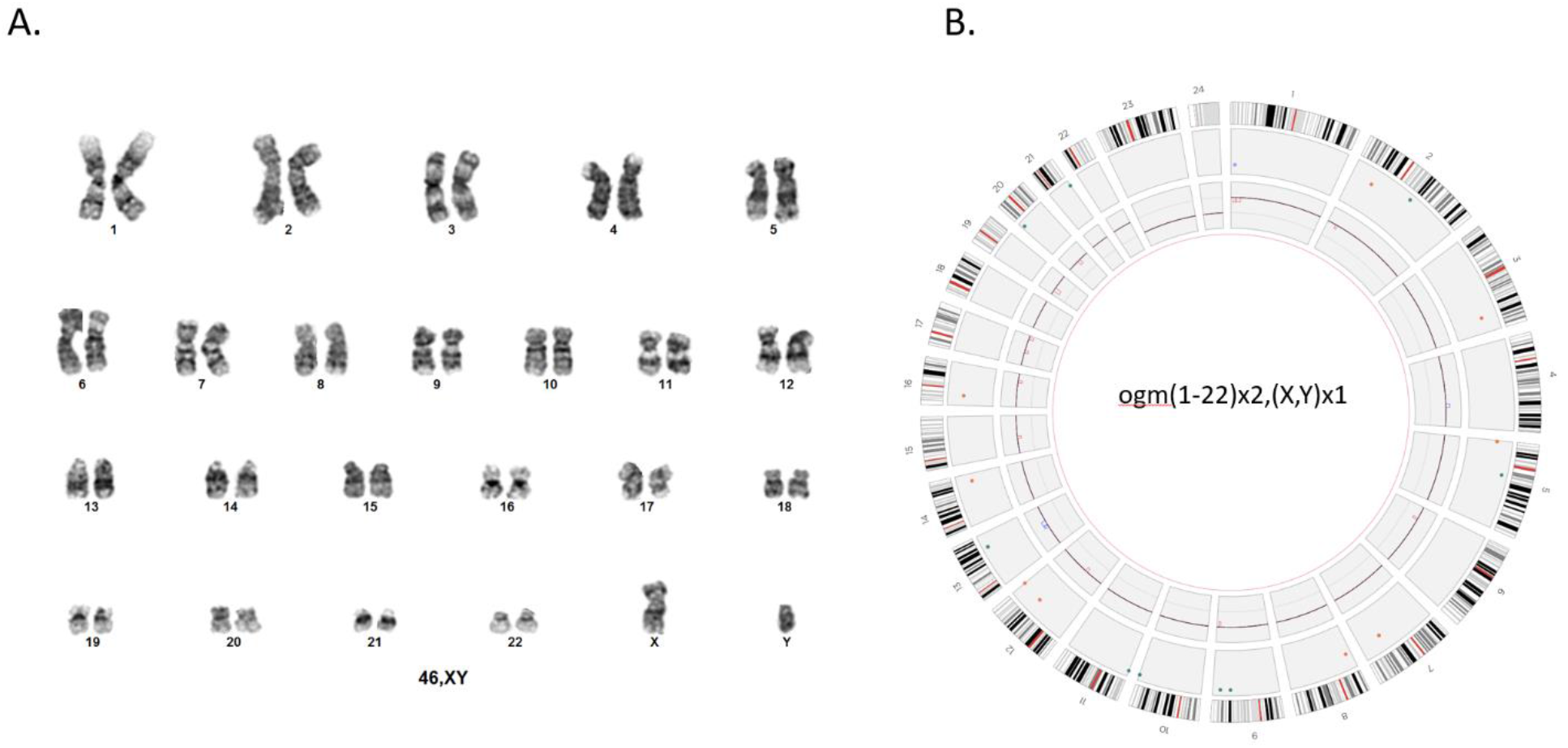
Karyotype and Optical Genome Mapping. **A**. Representation of the normal human male chromosomes (46,XY) with G-banded karyotype. **B**. Circos plot representation of a normal OMG profile of a male individual. The OGM nomenclature in this case is represented by ogm(1-22)x2,(X,Y)x1. The circos plot generated by Bionano Access is composed of 3 layers. The outer layer represents in silico locations of G-bands of each chromosome. The middle layer indicates the locations of the SVs that were identified in each chromosome. The inner layer represents the copy number changes throughout the chromosomes. In this particular case chromosomes 1-22 are at 2x and X,Y are at 1x. Note: chromosome X=23, and Y=24.

**Supplementary Figure 2:**
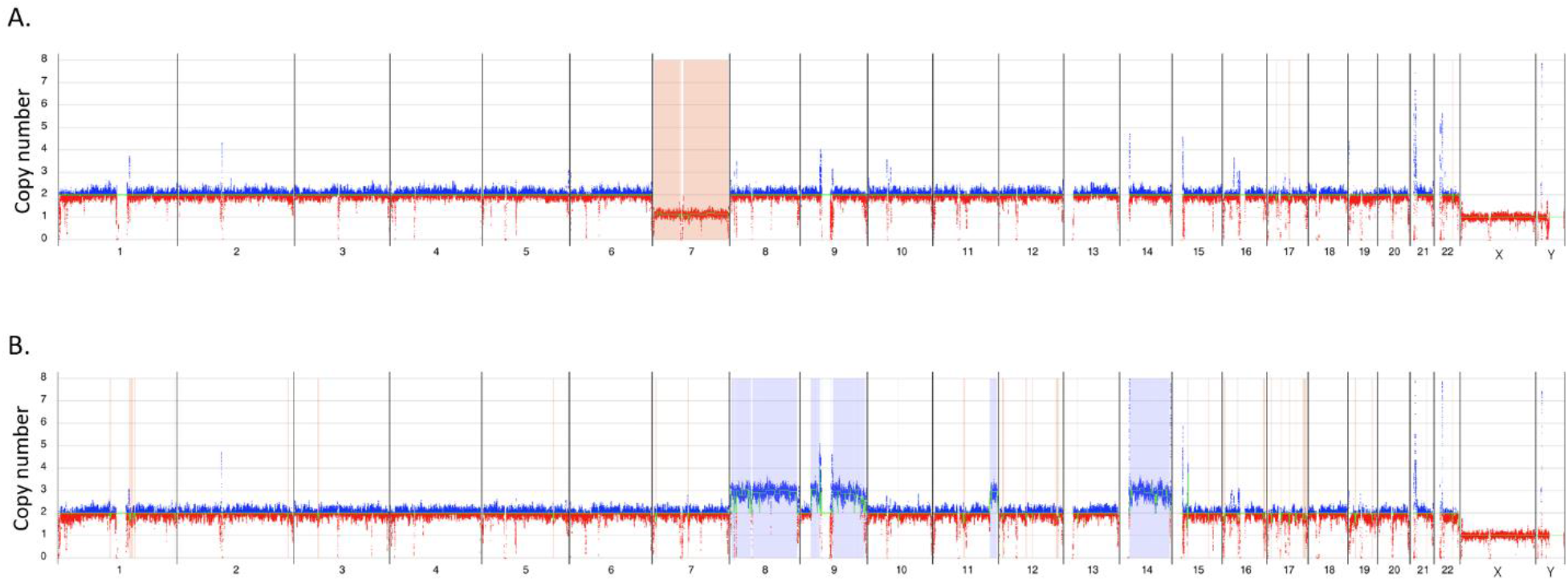
Classic AML copy number changes detected by Optical Genome Mapping. Whole genome CNV profiles: Y axis shows copy number range from 0-8 for each of the chromosomes – X axis. Molecules showing regions with increased copy number from the baseline are shown in blue and regions with decreased copy number are shown in red. **A**. Loss of chromosome 7, sample 83. **B**. Triplications of chromosomes 8, 9 and 14, sample – 62.

**Supplementary Figure 3:**
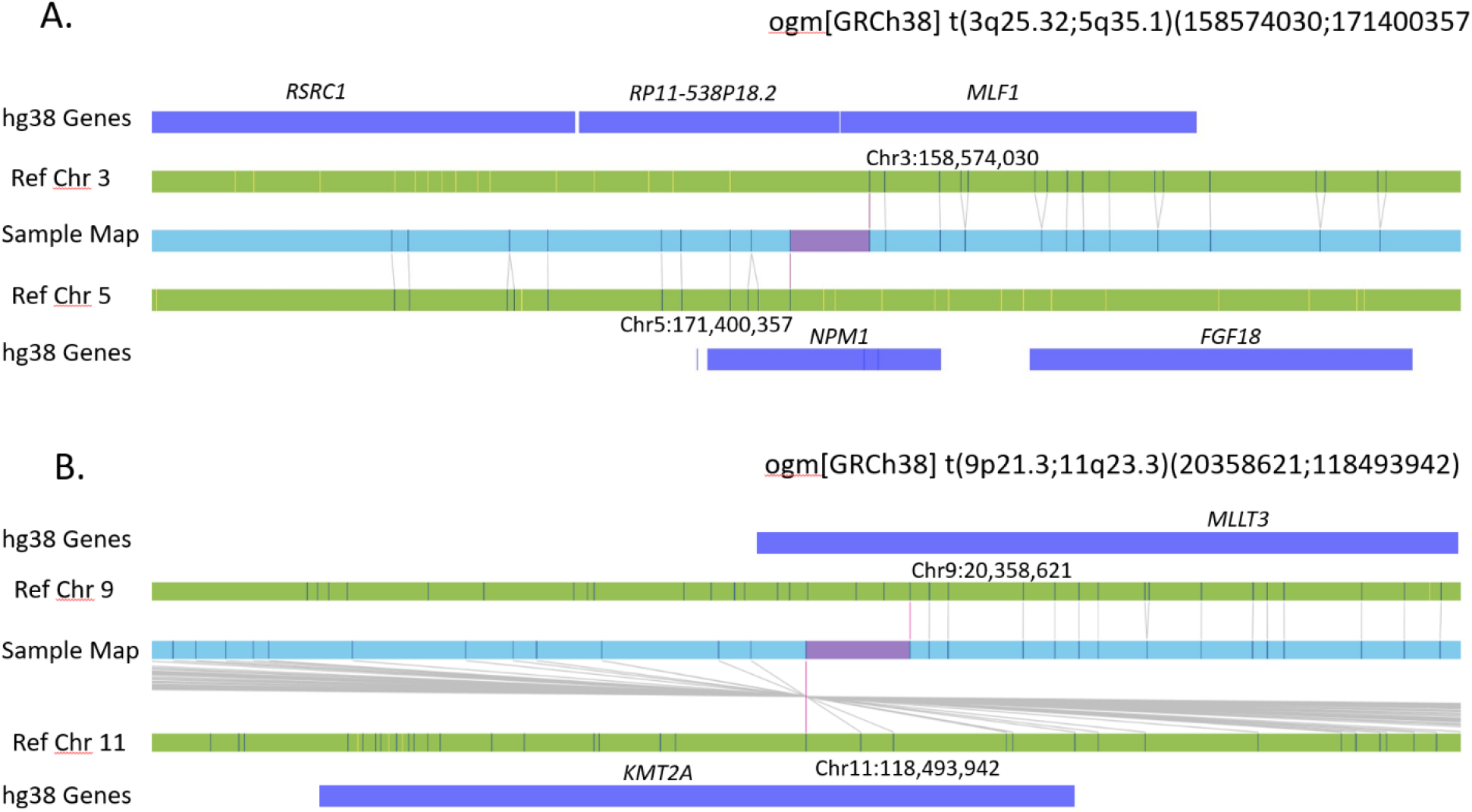
Classic AML translocations detected by Optical Genome Mapping. GRCh38 reference chromosomes with OGM label patterns are shown in green/black respectively. Assembled sample maps with label patters are shown in light blue. Label alignments between two maps are shown in grey strings. Translocation breakpoints are highlighted in purple. Overlapping genes are shown in blue. **A**. Translocation between chromosome 3 and 5, overlapping genes *MLF1* and *NPM1* seen in samples 67,68. **B**. Translocation between chromosome 9 and 11, overlapping genes *MLLT3* and *KMT2A* seen in samples 78,79.

